# Risk-based vaccination reveals marked heterogeneity in the clinical benefit of PCV20

**DOI:** 10.64898/2026.08.31.26361811

**Authors:** Hila Markovits, Yarden Jordi Cohen, Daniel Grupel, Ruth Goldstein, Hagit Goldenstein, Naama Katz Hanein, T. Razi, Yochai Schonmann, R. Arbel, Doron Netzer, Sophia Eilat Tsanani, Dan Yamin

## Abstract

Pneumococcal vaccination of older adults is primarily guided by age and clinical eligibility, despite substantial variation in individual risk of severe pneumonia. Here, we used longitudinal electronic health records from 787,538 adults aged ≥65 years to evaluate the real-world effectiveness of the 20-valent pneumococcal conjugate vaccine (PCV20) and quantify clinical benefit according to baseline risk of pneumonia hospitalization. We developed and validated a machine-learning model using pre-PCV20 data to estimate individual 12-month hospitalization risk and integrated these predictions into a propensity score matching framework. Overall vaccine effectiveness against pneumonia hospitalization was 16.5% (95% CI, 10.6– 22.1), but this population-level estimate masked substantial heterogeneity in clinical benefit. The 60% at lowest predicted risk, characterized by younger age and fewer pulmonary and other chronic conditions, showed no measurable reduction in hospitalization (VE, 3.1%; 95% CI, −14.4 to 18.0) and had an estimated 1-year number needed to vaccinate (NNV) of 7,423, compared with 184 and 115 in the intermediate- and high-risk groups, respectively. These findings suggest that incorporating baseline risk into adult pneumococcal vaccination strategies could enable more targeted and potentially better-timed vaccination.

## Introduction

Streptococcus pneumoniae is a leading cause of community-acquired pneumonia and invasive infections, including bacteremia and meningitis, and remains a major cause of morbidity and health-care utilization worldwide. The burden of pneumococcal disease is greatest among older adults and individuals with chronic medical conditions or immunocompromising disorders. ^1–3^

Pneumococcal conjugate vaccination has substantially reduced the burden of pneumococcal disease in older adults. In the CAPiTA randomized trial, the 13-valent pneumococcal conjugate vaccine (PCV13) reduced vaccine-type community-acquired pneumonia and invasive pneumococcal disease among adults aged 65 years and older, although no efficacy was demonstrated against all-cause pneumonia. Subsequent real-world studies supported the effectiveness of PCV13 against pneumococcal disease and reported reductions in pneumonia-related hospitalizations among older adults. ^4–6^

Despite widespread PCV13 use, a substantial burden of community-acquired pneumonia persists, with an increasing proportion attributable to serotypes not included in PCV13. ^7,8^ These residual serotypes account for a substantial proportion of invasive pneumococcal disease and community-acquired pneumonia in older adults and provided the rationale for the development of higher-valency conjugate vaccines. Moreover, the burden of severe pneumonia is highly heterogeneous among older adults ^9^ suggesting that the absolute benefit of vaccination may differ substantially according to baseline risk. PCV20 was therefore developed by incorporating seven additional serotypes associated with persistent pneumococcal disease burden beyond those covered by PCV13. ^10,11^

Unlike PCV13, whose recommendations were supported by randomized clinical endpoint data, evidence supporting PCV20 in adults was initially derived from immunogenicity and safety studies. Regulatory approval and subsequent vaccination recommendations were based on immunobridging studies demonstrating non-inferior immune responses compared with PCV13 and generally favorable comparisons with the 23-valent pneumococcal polysaccharide vaccine (PPSV23) ^10–14^ These studies did not evaluate clinical outcomes such as pneumonia-related hospitalization. A recent large real-world effectiveness study demonstrated protection against invasive pneumococcal disease, pneumococcal pneumonia, all-cause pneumonia, and lower respiratory tract infection.^15^ However, pneumonia-related hospitalization was not evaluated, and individuals with recent PCV13 or PPSV23 vaccination were largely excluded. In addition, the extent to which the clinical benefit of PCV20 varies according to baseline risk remains unknown.

To address these gaps, we evaluated the real-world effectiveness of PCV20 against pneumonia-related hospitalization among adults 65 years of age or older. We also quantified the absolute clinical benefit of vaccination across the spectrum of baseline risk in a large population-based cohort using a risk-informed propensity score-matching approach.

## Methods

### Study design

We conducted a retrospective cohort study using longitudinal electronic health record data from Clalit Health Services (CHS) to estimate the real-world effectiveness of the 20-valent pneumococcal conjugate vaccine (PCV20) against pneumonia-related hospitalization among adults aged 65 years or older.

CHS is the largest integrated health-care provider in Israel serving approximately 5 million members ^16^, representing about 50.6% of the Israeli population ^16–18^. CHS members are broadly representative of the Israeli population and encompass all major demographic, ethnic, and socioeconomic groups. Clinical information is routinely collected during health-care encounters and stored in a centralized electronic health record system. The database includes demographic characteristics, diagnoses, hospitalizations, outpatient visits, laboratory test results, medication dispensing, and vaccinations ^18^. Data are coded, pseudonymized, stored, and analyzed within the secure CHS research platform, which is accessible only to authorized researchers. We accessed individual-level information on demographic characteristics, vaccination history, comorbid conditions, health-care utilization, and clinical outcomes.

The study period extended from June 1, 2023, to July 31, 2025, covering two consecutive respiratory seasons. Members of the study cohort were assessed monthly to identify individuals who had reached 65 years of age and became eligible for inclusion. Participants were followed from the beginning of each season until the end of the corresponding respiratory season. Baseline characteristics and comorbidities were assessed before the index date using available medical history within the CHS database.

The institutional review board approved the study and waived the requirement for informed consent because the analysis used de-identified routinely collected health care data (protocol number 0139-21-CHS).

### Participants

Eligible participants were adults aged 65 years or older who were members of Clalit Health Services (CHS) during the study period. To be eligible in a given month, individuals were required to be alive at the beginning of the month, continuously enrolled in CHS throughout the corresponding calendar year, and to have had at least one recorded health-care interaction, including a vaccination, outpatient visit, laboratory test, hospitalization, or medication dispensing event, during the preceding year.

To reduce bias arising from differences in health-care utilization and vaccine-seeking behavior, we excluded individuals with neither a documented history of pneumococcal vaccination, including 13-valent pneumococcal conjugate vaccine (PCV13), 20-valent pneumococcal conjugate vaccine (PCV20), or 23-valent pneumococcal polysaccharide vaccine (PPSV23), nor a recorded influenza vaccination during the preceding three years ^16,19^. Individuals meeting neither criterion represent a distinct population with persistently low uptake of preventive health services and adult vaccinations and are unlikely to receive PCV20 during routine clinical care. Restricting the analysis to individuals with evidence of prior participation in vaccination programs reduced the potential for residual confounding related to vaccine-seeking behavior while preserving the representativeness of the study population. Notably, more than 92% of CHS members aged 65 years or older met this criterion and were retained in the analysis.

### Exposure definition

The exposure of interest was receipt of the 20-valent pneumococcal conjugate vaccine (PCV20), as recorded in the CHS vaccination registry. Eligibility and exposure status were assessed monthly. In each calendar month, individuals who had received PCV20 at least 30 days earlier and were eligible for analysis contributed to the vaccinated group, whereas eligible individuals who had not yet received PCV20 were eligible to serve as matched unvaccinated controls. To allow for the development of vaccine-induced immunity and avoid exposure misclassification, participants were excluded from both the vaccinated and unvaccinated groups during the 30 days immediately after vaccination.

### Outcome measures

The outcome was pneumonia-related hospitalization. Pneumonia-related hospitalization was defined as an inpatient hospitalization with a diagnosis of pneumonia. The outcome definition was prespecified and based on ICD-9-CM diagnosis codes (Appendix Table S2).

### Statistical analysis

#### Sequential risk-stratified propensity score matching

Because baseline pneumonia risk varies substantially among older adults, we sought to balance participants according to both their predicted risk of pneumonia hospitalization and their probability of receiving PCV20. We therefore developed a sequential risk-stratified propensity score–matching approach in which participants were first stratified according to their predicted 12-month risk of pneumonia hospitalization and then according to their propensity to receive PCV20 before matching.

A pneumonia risk prediction model was developed using extreme gradient boosting (XGBoost). The model was trained on CHS data from 2017 through 2021 and evaluated on data from 2022 to predict each participant’s 12-month risk of pneumonia hospitalization. Model development was restricted to the pre-PCV20 period to avoid incorporating information from PCV20-vaccinated individuals. Candidate predictors included demographic characteristics, comorbidities, community- or hospital-diagnosed pneumonia in the previous year^20^, all-cause hospitalizations, vaccination history, health care utilization, and other clinical variables (Appendix Table S1).

At the beginning of each respiratory season, the model was applied using each participant’s current clinical characteristics to estimate their predicted 12-month risk of pneumonia hospitalization. Participants were ranked according to their predicted risk and partitioned into 25 strata with approximately equal cumulative predicted risk, corresponding to similar expected numbers of pneumonia hospitalizations in each stratum.

A second XGBoost model was developed using the 2023-2025 study data to estimate each participant’s propensity to receive PCV20. Participants were classified according to age group (65-69, 70-79, and ≥80 years), predicted pneumonia risk (25 strata), and propensity to receive PCV20 (10 strata), yielding 750 age-risk-propensity strata. Matching was performed monthly throughout each respiratory season by randomly matching PCV20-vaccinated participants to eligible unvaccinated controls within the same calendar month and age-risk-propensity stratum using bootstrap resampling. This approach allowed newly eligible participants and newly vaccinated participants to enter the analysis over time.

### Vaccine effectiveness estimation

For each bootstrap iteration, monthly matching was performed throughout the respiratory season. Outcome events were accumulated over the entire respiratory season, and relative risks (RRs) were estimated by comparing the cumulative risk of pneumonia hospitalization between vaccinated and matched unvaccinated participants. Vaccine effectiveness (VE) was calculated as (1 - RR) × 100%. Analyses were performed separately for the 2023-2024 and 2024-2025 respiratory seasons and after pooling both seasons. Overall VE was estimated by pooling the matched cohorts before calculating the RR. The complete bootstrap matching procedure was repeated 1,000 times. Point estimates were calculated as the mean across bootstrap iterations. Ninety-five percent confidence intervals were obtained using the non-parametric percentile bootstrap method from the 2.5th and 97.5th percentiles of the bootstrap distribution.

Subgroup analyses were performed using the same matching procedure after stratification by age group, predicted risk category, and presence of an immunocompromising condition. As a post hoc analysis, the 25 predicted risk strata were consolidated into three clinically interpretable risk categories to facilitate subgroup analyses. The low-, medium-, and high-risk categories comprised approximately 60%, 30%, and 10% of the study population, respectively, corresponding to participants with the lowest, intermediate, and highest predicted risks of pneumonia hospitalization. Immunocompromising conditions were identified using the Clalit Health Services chronic disease registry ^21^, which integrates diagnoses, medication records, laboratory results, and other clinical information through validated automated algorithms. Immunocompromising conditions included solid organ transplantation, dialysis-dependent kidney disease, and anatomic or functional asplenia.^22^

Subgroup analyses were performed using the same bootstrap matching procedure after stratification by age (65-69, 70-79, and ≥80 years), predicted baseline pneumonia risk, previous pneumococcal vaccination history (prior PCV13, prior PPSV23, or neither), and the presence of an immunocompromising condition. For presentation of subgroup analyses, the 25 predicted risk strata were consolidated post hoc into three clinically interpretable categories corresponding to the 0th–60th, 60th–90th, and 90th–100th percentiles of predicted pneumonia risk, based on the distribution of predicted risk in the pre-PCV20 development cohort (2017-2021) and validated in 2022.

Vaccine effectiveness was estimated separately within each subgroup using the same bootstrap matching procedure. Yearly outcome risks were estimated from the matched cohorts. The 1-year absolute risk reduction was calculated as the difference in yearly risk between vaccinated and matched unvaccinated participants, and the corresponding number needed to vaccinate (NNV) was estimated as the reciprocal of the absolute risk reduction, both overall and within prespecified subgroups.

An alternative analysis using conventional covariate-based matching was performed to evaluate the robustness of the primary findings ^19,23^ (Appendix Section 3). All analyses were performed using Python version 3.9.7.

## Results

### Study population and baseline characteristics

Of the 787,538 potentially eligible CHS members, 139,540 and 184,256 participants were excluded before matching in the 2023–2024 and 2024–2025 respiratory seasons, respectively (Figure 1). After applying the eligibility criteria, 647,998 participants were eligible for matching in 2023–2024 and 603,282 in 2024–2025. During the respective seasons, 89,764 and 294,753 participants were eligible for analysis as PCV20-vaccinated, whereas 647,947 and 487,160 participants were eligible as unvaccinated.

**Figure 1.**
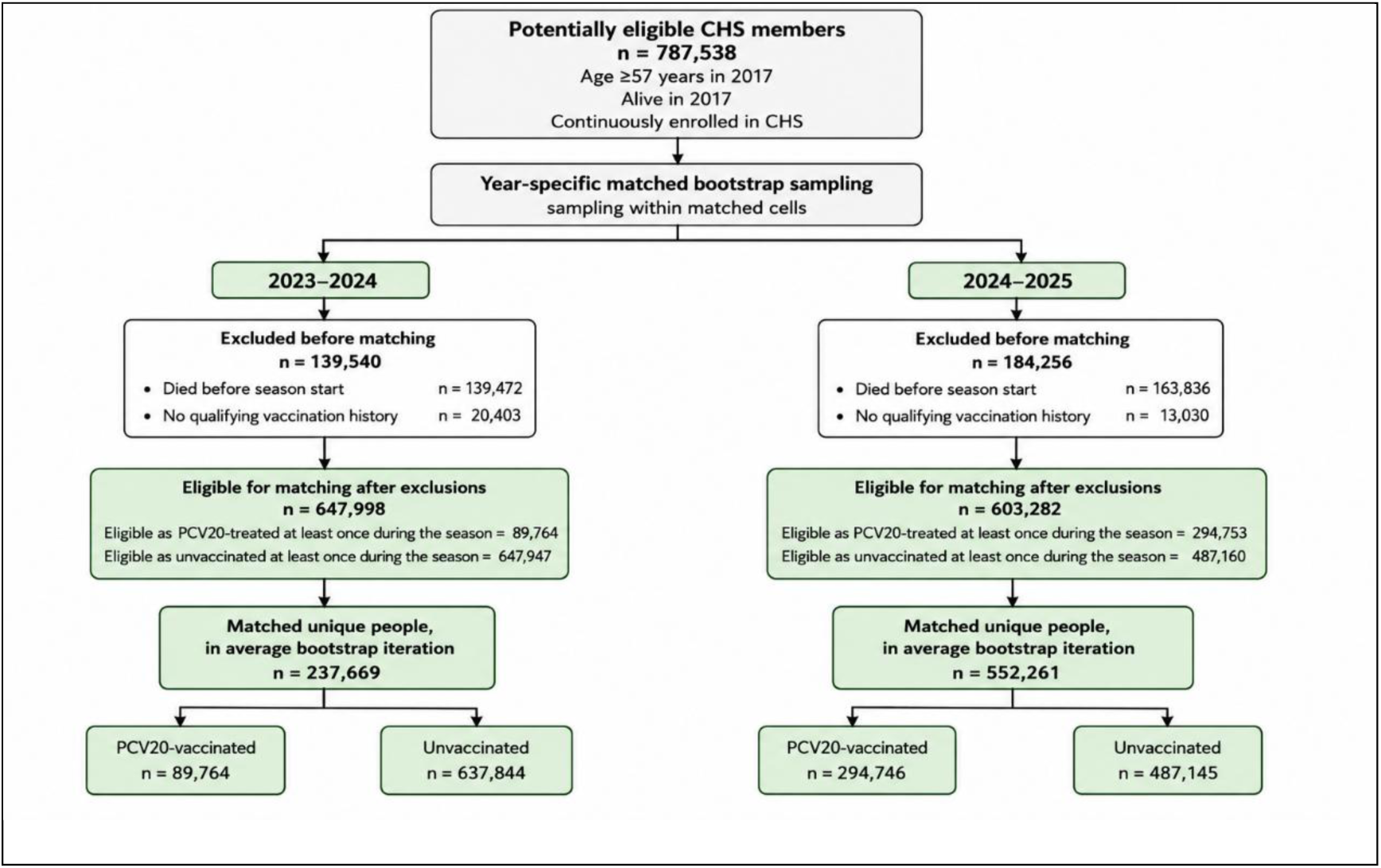
**Study profile**

The uptake of pneumococcal vaccines during the study period and the distribution of previous pneumococcal vaccination histories are shown in the Appendix (Figure S1). Baseline characteristics of vaccinated participants and matched unvaccinated controls are summarized in Table 1. Following sequential risk-stratified propensity score matching, baseline characteristics were well balanced, with standardized mean differences below 0.1 for all prespecified covariates. Matching achieved exact balance across age groups and the 25 predicted pneumonia risk strata (all standardized mean differences = 0.000); for presentation, the latter are shown in Table 1 as three clinically interpretable percentile-based risk categories. As expected, the underlying 25 risk strata showed progressively increasing observed rates of pneumonia hospitalization (Appendix Figure S2).

**Table 1.** Baseline Characteristics of PCV20-Vaccinated Participants and Matched Unvaccinated Controls by Respiratory Season.

| Characteristic | Category | 2023 PCV20-treated<br>(n = 89,764) | 2023 Matched Controls<br>(n = 637,844) | 2023<br>SMD | 2024 PCV20-treated<br>(n = 294,746) | 2024 Matched Controls<br>(n = 487,145) | 2024<br>SMD |
| --- | --- | --- | --- | --- | --- | --- | --- |
| Age group | 65–69 | 21.1% (20.9–21.2) | 21.1% (20.9–21.2) | 0.000 | 21.8% (21.7–21.9) | 21.8% (21.7–21.9) | 0.000 |
|  | 70–79 | 59.0% (58.8–59.2) | 59.0% (58.8–59.2) |  | 57.6% (57.5–57.7) | 57.6% (57.5–57.7) |  |
|  | ≥80 | 20.0% (19.8–20.1) | 20.0% (19.8–20.1) |  | 20.6% (20.5–20.7) | 20.6% (20.5–20.7) |  |
| Sex | Female | 53.0% (52.8–53.2) | 55.3% (55.1–55.5) | 0.047 | 53.3% (53.2–53.4) | 56.2% (56.1–56.3) | 0.059 |
|  | Male | 47.0% (46.8–47.3) | 44.7% (44.5–44.9) |  | 46.7% (46.6–46.8) | 43.8% (43.7–43.9) |  |
| Predicted pneumonia risk* | Low (0th–60th percentile) | 60.3% (60.0–60.5) | 60.3% (60.0–60.5) | 0.000 | 58.6% (58.5–58.7) | 58.6% (58.5–58.7) | 0.000 |
|  | Medium (60th–90th percentile) | 30.7% (30.5–30.9) | 30.7% (30.5–30.9) |  | 31.3% (31.2–31.4) | 31.3% (31.2–31.4) |  |
|  | High (90th–100th percentile) | 9.0% (8.9–9.1) | 9.0% (8.9–9.1) |  | 10.2% (10.1–10.2) | 10.2% (10.1–10.2) |  |
| Previous pneumococcal vaccination† | Previous PCV13 | 18.2% (18.1–18.4) | 16.2% (16.0–16.3) | 0.055 | 13.2% (13.2–13.3) | 12.5% (12.5–12.6) | 0.021 |
|  | Previous PPSV23 | 28.4% (28.2–28.6) | 32.3% (32.1–32.5) | 0.086 | 29.3% (29.2–29.4) | 30.3% (30.2–30.4) | 0.021 |
|  | No previous PCV13/PPSV23 | 57.1% (56.9–57.3) | 55.1% (54.9–55.3) | 0.040 | 60.3% (60.2–60.4) | 60.1% (60.0–60.2) | 0.005 |
| Health-care utilization | ≥1 hospitalization during previous year‡ | 17.7% (17.5–17.9) | 17.5% (17.3–17.7) | 0.006 | 17.5% (17.4–17.5) | 17.1% (17.0–17.2) | 0.010 |
|  | ≥1 hospitalization 1–3 years before baseline‡ | 27.7% (27.5–27.9) | 28.4% (28.2–28.6) | 0.016 | 28.6% (28.5–28.7) | 28.8% (28.8–28.9) | 0.005 |
| **Pneumonia history** | Pneumonia during previous year | 1.2% (1.2–1.3) | 1.1% (1.0–1.1) | 0.014 | 1.2% (1.2–1.2) | 1.2% (1.2–1.2) | 0.002 |
|  | Pneumonia 1–3 years before baseline only | 1.5% (1.5–1.6) | 1.5% (1.5–1.6) | 0.002 | 1.6% (1.6–1.6) | 1.6% (1.6–1.7) | 0.005 |
| **Immunodeficiency** | Yes | 7.5% (7.4–7.6) | 6.3% (6.2–6.4) | 0.045 | 6.8% (6.8–6.9) | 6.3% (6.3–6.3) | 0.022 |
| **Diabetes** |  | 35.8% (35.6–36.0) | 36.9% (36.7–37.1) | 0.023 | 38.4% (38.3–38.5) | 37.3% (37.2–37.4) | 0.022 |
| **Liver disease** |  | 2.4% (2.3–2.5) | 2.3% (2.3–2.4) | 0.004 | 2.5% (2.5–2.5) | 2.3% (2.3–2.4) | 0.010 |
| **Malignancy** |  | 24.1% (23.9–24.3) | 21.0% (20.8–21.1) | 0.076 | 22.6% (22.5–22.6) | 21.1% (21.0–21.2) | 0.035 |
| **Neurologic disease** |  | 4.4% (4.3–4.5) | 5.2% (5.1–5.3) | 0.040 | 5.6% (5.5–5.6) | 6.7% (6.7–6.8) | 0.047 |
| **Obesity** |  | 38.7% (38.5–39.0) | 39.2% (39.0–39.4) | 0.009 | 41.0% (40.9–41.1) | 39.6% (39.5–39.7) | 0.030 |
| **Chronic pulmonary disease** |  | 15.5% (15.3–15.6) | 13.1% (12.9–13.2) | 0.068 | 14.5% (14.4–14.6) | 13.0% (12.9–13.0) | 0.045 |
| **Smoking history§** | Current or former | 44.4% (44.1–44.6) | 42.2% (42.0–42.3) | 0.039 | 43.7% (43.6–43.8) | 42.2% (42.1–42.3) | 0.030 |

\* Risk categories were derived from the XGBoost prediction model developed using pre-PCV20 data (2017–2021) and validated in 2022. Categories correspond to the 0th–60th, 60th–90th, and 90th–100th percentiles of predicted risk.
† Categories are not mutually exclusive; participants may have received both PCV13 and PPSV23.
‡ Indicates at least one hospitalization during the specified interval before baseline.
§ Smoking history includes current and former smokers.
Values are percentages (95% bootstrap confidence intervals) unless otherwise indicated.

### Effectiveness of PCV20 and clinical impact

Estimated vaccine effectiveness was consistent across the 2023-2024 and 2024-2025 respiratory seasons (Figure 2). Vaccine effectiveness against pneumonia hospitalization was 16.1% (95% CI, −0.4 to 30.6) and 16.5% (95% CI, 9.7 to 22.6), respectively, yielding a pooled estimate of 16.5% (95% CI, 10.6 to 22.1). Annual outcome risks are shown in Table 2. A sensitivity analysis using conventional covariate-based matching yielded similar estimates (Appendix Section 3).

**Figure 2.**
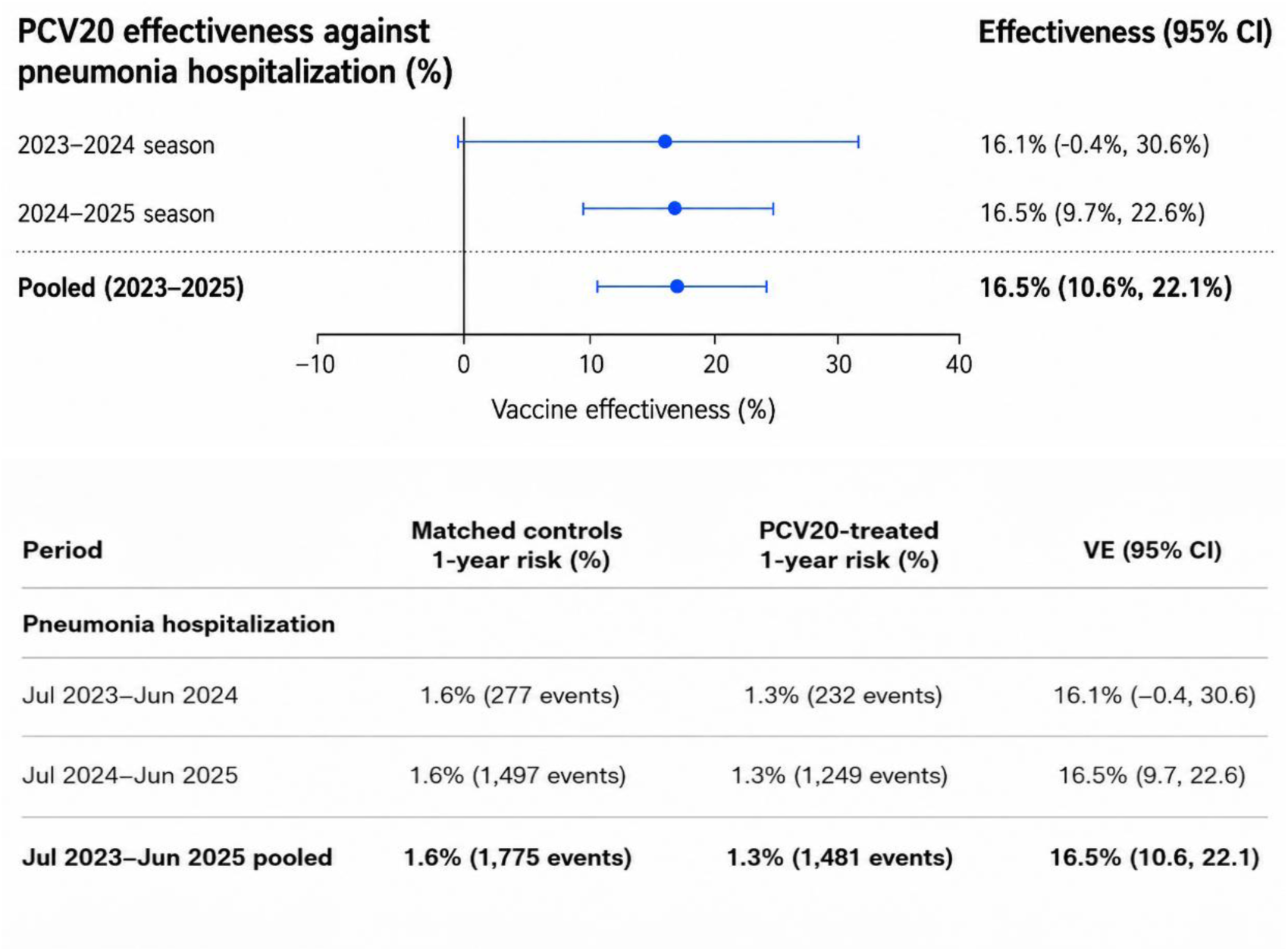
Year-specific and pooled effectiveness of PCV20 against pneumonia hospitalization. **(A)** Forest plots showing vaccine effectiveness estimates for each respiratory season and for the pooled study period. Points represent vaccine effectiveness estimates, and horizontal lines represent 95% confidence intervals. **(B)** Corresponding annualized 1-year risks among PCV20-vaccinated participants and matched unvaccinated controls. The mean number of outcome events across bootstrap iterations is shown in parentheses. Pooled estimates combine the July 2023 to June 2024 and July 2024 to June 2025 respiratory seasons. VE = vaccine effectiveness; CI = confidence interval.

**Table 2.** Absolute Clinical Benefit of PCV20 across Prespecified Subgroups.

| Subgroup | Level | PCV20-treated 1-year risk (%) | Matched controls 1-year risk (%) | Absolute Risk Reduction (%) | NNV (95% CI) |
| --- | --- | --- | --- | --- | --- |
| **Overall** | — | **1.3 (1.3–1.4)** | **1.6 (1.5–1.7)** | **0.26 (0.16–0.36)** | **382 (278–623)** |
| **Age** | 65–69 years | 0.7 (0.6–0.8) | 0.8 (0.7–0.9) | 0.09 (–0.06–0.24) | 1164 (CI includes ∞) |
|  | 70–79 years | 1.0 (0.9–1.1) | 1.3 (1.2–1.4) | 0.25 (0.13–0.37) | 397 (270–744) |
|  | ≥80 years | 2.8 (2.6–3.1) | 3.3 (3.1–3.5) | 0.47 (0.17–0.77) | 212 (129–598) |
| **Predicted pneumonia risk\*** | Low (0th–60th percentile) | 0.4 (0.4–0.5) | 0.4 (0.4–0.5) | 0.01 (–0.06–0.09) | **7423 (CI includes ∞)** |
|  | Medium (60th–90th percentile) | 1.6 (1.4–1.7) | 2.1 (2.0–2.3) | 0.54 (0.35–0.75) | **184 (134–286)** |
|  | High (90th–100th percentile) | 6.8 (6.4–7.3) | 7.7 (7.2–8.3) | 0.87 (0.09–1.58) | **115 (63–1067)** |
| **Previous pneumococcal vaccination** | Prior PCV13 | 1.3 (1.0–1.5) | 1.5 (1.2–1.9) | 0.29 (–0.11–0.70) | 342 (CI includes ∞) |
|  | Prior PPSV23 | 0.8 (0.6–0.9) | 0.9 (0.8–1.1) | 0.14 (–0.07–0.35) | 721 (CI includes ∞) |
|  | No prior PCV13/PPSV23 | 1.7 (1.6–1.8) | 2.0 (1.9–2.2) | 0.31 (0.13–0.49) | 325 (205–746) |
| **Immunocompromising condition** |  | 4.0 (2.3–5.8) | 4.4 (2.7–6.3) | 0.41 (–2.26–2.98) | 246 (CI includes ∞) |

Abbreviations: NNV, number needed to vaccinate.
Values are annual risks (%) with 95% bootstrap confidence intervals unless otherwise indicated.
\* Predicted pneumonia risk categories correspond to the 0th–60th, 60th–90th, and 90th–100th percentiles of predicted 12-month pneumonia hospitalization risk derived from the pre-PCV20 prediction model. These corresponded to observed annual pneumonia hospitalization risks among matched unvaccinated controls of approximately 0.0–0.7%, 0.8–2.1%, and 2.2–7.7%, respectively.

To examine whether the clinical benefit of PCV20 varied according to baseline risk, we grouped participants into three categories corresponding to the 0th–60th, 60th– 90th and 90th–100th percentiles of predicted pneumonia hospitalization risk (Fig. 3), comprising approximately 60%, 30% and 10% of the study population, respectively. To facilitate clinical interpretation, we derived a simplified score based on the characteristics most strongly associated with predicted risk (Fig. 3). The score assigned one point for age 75–79 years, two points for age ≥80 years, and one point each for chronic pulmonary disease, respiratory hospitalization within the previous 3 years and the presence of ≥2 active chronic comorbidities. Scores of 0–1, 2–3 and 4–5 points provided a clinical approximation of the low-, medium- and high-risk groups, respectively. Additional interpretation of the prediction model is provided in Extended Data Fig. S3. PCV20 effectiveness differed across predicted risk groups. Among the 60% of older adults classified as low risk, no measurable reduction in pneumonia hospitalization was observed (VE, 3.1%; 95% CI, −14.4 to 18.0). By contrast, vaccination was associated with reduced pneumonia hospitalization in the medium- and high-risk groups (Fig. 4). Analyses stratified by age and previous pneumococcal vaccination history generally favored vaccination, although confidence intervals crossed zero in several smaller subgroups, including adults aged 65–69 years and those previously vaccinated with PCV13 or PPSV23 (Fig. 4).

**Figure 3.**
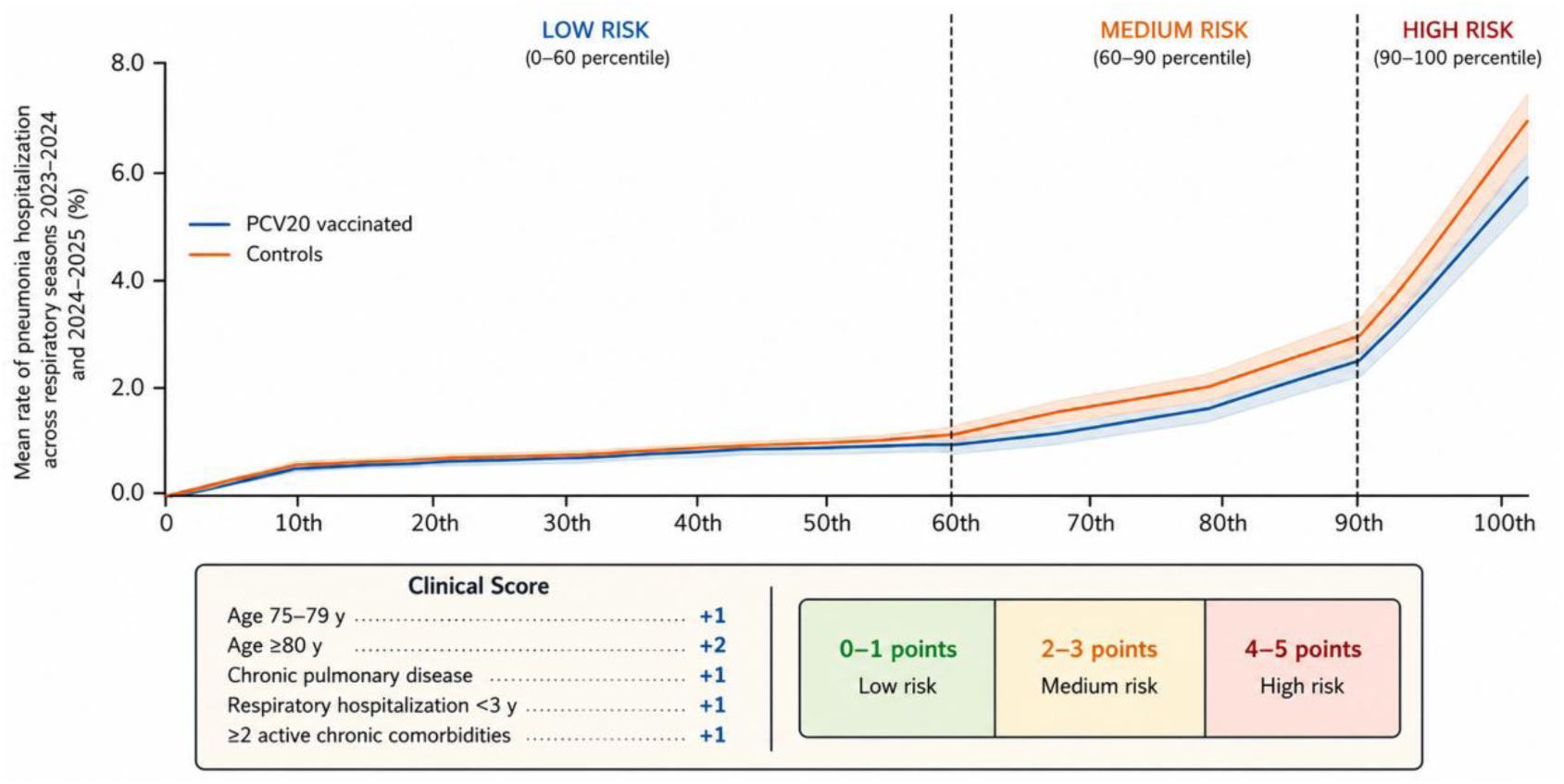
Annual Pneumonia Hospitalization Rates among PCV20-Vaccinated Participants and Matched Unvaccinated Controls across the Predicted Risk Distribution. Annual pneumonia hospitalization rates among PCV20-vaccinated participants and matched unvaccinated controls are shown across the distribution of predicted baseline pneumonia risk. Risk groups correspond to the 0th-60th, 60th-90th, and 90th-100th percentiles of predicted risk derived from the pre-PCV20 prediction model. Shaded areas indicate 95% confidence intervals. The inset presents a simplified clinical approximation of the XGBoost-derived risk groups based on age, chronic pulmonary disease, previous respiratory hospitalization, and multimorbidity. Additional interpretation of the prediction model is provided in Appendix Figure S3A. The four-level surrogate decision tree that informed this clinical approximation is shown in Appendix Figure S3B and was used for clinical interpretation only.

**Figure 4.**
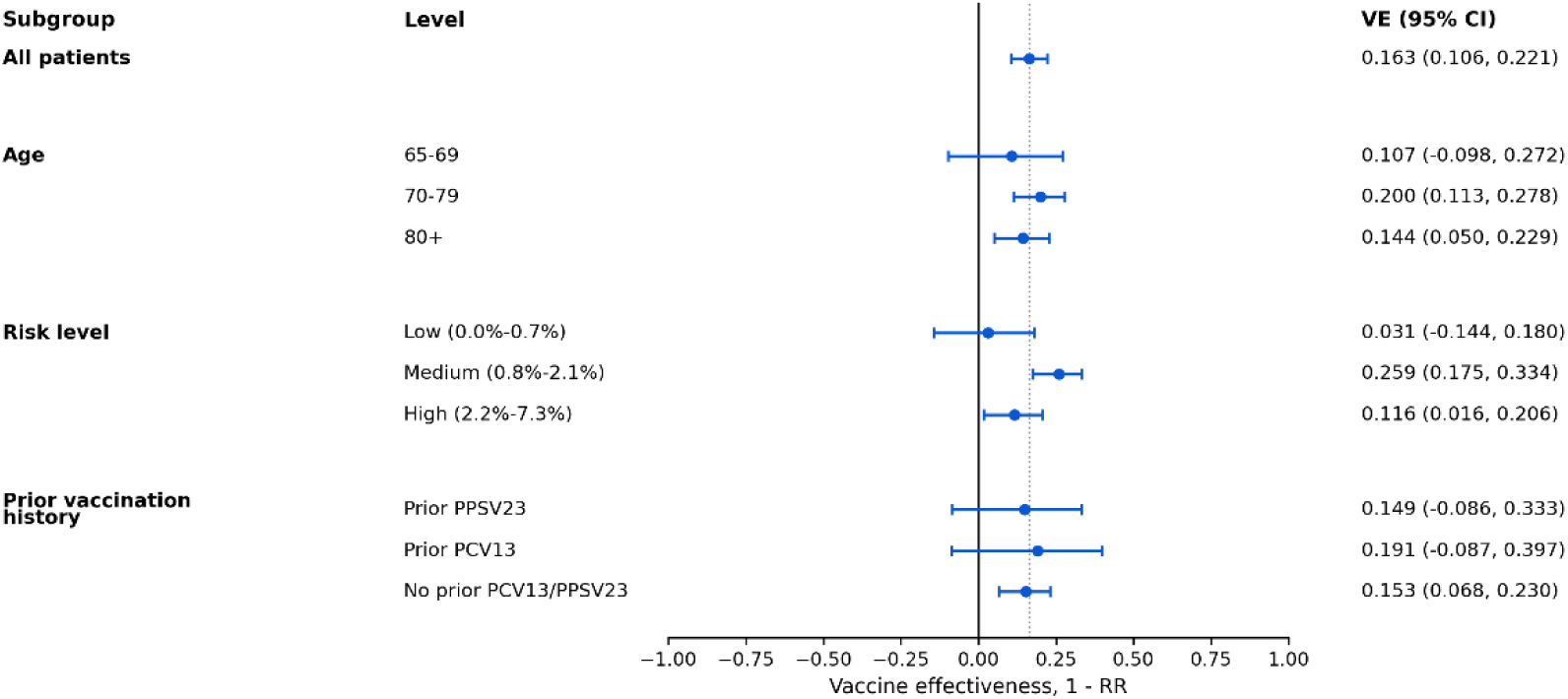
Subgroup analyses of PCV20 effectiveness against pneumonia hospitalization. * Risk groups were based on XGBoost-derived predicted pneumonia hospitalization risk, with participants categorized into low-, medium-, and high-risk groups before outcome assessment. We present here the approximate clinical score for clarity. The full surrogate decision tree used for clinical interpretation is provided in Appendix Figure S3B.

Differences were more pronounced when vaccination benefit was considered on an absolute scale (Table 2). The estimated 1-year number needed to vaccinate (NNV) was 7,423 in the low-risk group, compared with 184 in the medium-risk group and 115 in the high-risk group, representing an approximately 65-fold difference between the lowest- and highest-risk groups. A similar gradient was observed across age groups, with the NNV decreasing from 1,164 among adults aged 65–69 years to 212 among those aged ≥80 years. Absolute clinical benefit was also greater among immunocompromised participants, although estimates were imprecise owing to the smaller sample size. Thus, the overall effectiveness of PCV20 masked substantial differences in clinical benefit according to baseline risk.

## Discussion

In this large population-based study spanning two respiratory seasons, PCV20 vaccination was associated with a reduced risk of pneumonia hospitalization among adults aged 65 years or older, but its clinical benefit varied markedly according to baseline risk. Although the overall vaccine effectiveness was 16.5%, this population-level estimate masked substantial heterogeneity: among the 60% of older adults at lowest predicted risk, no measurable reduction in pneumonia hospitalization was observed, whereas vaccination was associated with substantially greater absolute benefit among individuals at medium and high risk. This translated into an approximately 65-fold difference in the number needed to vaccinate, from 7,423 in the low-risk group to 115 in the highest-risk group. These findings suggest that age-based eligibility alone may poorly distinguish individuals according to their expected clinical benefit from PCV20 and provide a rationale for evaluating risk-informed approaches to adult pneumococcal vaccination.

The distinction between these risk groups may have important implications for the timing of vaccination. The low-risk group, comprising approximately 60% of older adults, was characterized predominantly by younger age, absence of chronic pulmonary disease or recent respiratory hospitalization, and limited multimorbidity, and had both a very low baseline rate of pneumonia hospitalization and no measurable vaccine effectiveness. By contrast, individuals in the highest-risk group, characterized by advanced age, pulmonary disease, previous respiratory hospitalization and multimorbidity, had markedly higher baseline hospitalization rates and accounted for a disproportionate share of the absolute vaccine benefit. Consequently, estimates averaged across the entire older population are strongly influenced by this smaller high-risk group and may obscure the limited absolute benefit among the majority at low risk. Given that protection following pneumococcal vaccination may wane over time, vaccinating all individuals at the same age may not necessarily align vaccination with the period when an individual is most likely to benefit. Risk-informed vaccination could potentially allow vaccination to be better timed to increasing susceptibility to severe pneumonia.

Our findings complement the recently published Medicare-based evaluation of PCV20, the only previous real-world effectiveness study of this vaccine in adults. That study demonstrated protection against invasive pneumococcal disease, pneumococcal pneumonia, all-cause pneumonia, and lower respiratory tract infection. Although direct comparison is limited by differences in study design and outcome definitions, our findings extend those observations in several important ways. First, we evaluated a clinically meaningful severe outcome, specifically pneumonia-related hospitalization, rather than all-cause pneumonia irrespective of disease severity.

Second, our study included adults with recent previous PCV13 and PPSV23 vaccination, reflecting routine clinical practice and allowing evaluation of PCV20 in the context of sequential vaccination strategies. Finally, by accounting for baseline pneumonia risk and seasonal variation in respiratory disease, our analysis demonstrates how the clinical impact of PCV20 varies according to baseline risk in routine clinical practice.

The magnitude of vaccine effectiveness observed in our study is consistent with previous evaluations of PCV13 against all-cause pneumonia hospitalization in older adults. Retrospective cohort studies have reported modest effectiveness against this endpoint, with estimates ranging from approximately 7% to 10%. Hsiao and colleagues reported an adjusted vaccine effectiveness of 10.0% against hospitalization for all-cause pneumonia, whereas Kobayashi and colleagues estimated an overall effectiveness of 6.7%, with greater protection among individuals at lower baseline risk ^6,24^. Although the effectiveness estimate observed in our study was somewhat higher than those reported for PCV13, direct comparisons should be interpreted cautiously because of differences in vaccine valency, study populations, outcome definitions, and study design. Nevertheless, because pneumococci account for only a minority of all-cause pneumonia hospitalizations, a 16% reduction in this syndromic endpoint may correspond to a substantially larger effect against the vaccine-preventable pneumococcal fraction of disease. ^1^

By contrast, studies evaluating microbiologically confirmed pneumococcal pneumonia requiring hospitalization have consistently reported substantially higher vaccine effectiveness estimates. McLaughlin and colleagues reported vaccine effectiveness of 73% against hospitalized vaccine-type community-acquired pneumonia, whereas Heo and colleagues estimated approximately 40% effectiveness against hospitalized pneumococcal community-acquired pneumonia ^5,25^. These higher estimates are expected because microbiologically confirmed endpoints are more specific for vaccine-preventable pneumococcal disease than syndromic outcomes such as all-cause pneumonia. Additional differences in study design, study population, circulating serotypes, and outcome definitions are also likely to contribute.

This study has several limitations. First, pneumonia-related hospitalizations were identified using diagnosis codes rather than microbiological confirmation, precluding estimation of vaccine effectiveness against pneumococcal-specific disease or individual vaccine serotypes. Nevertheless, pneumonia-related hospitalization is a clinically meaningful outcome that directly reflects severe disease and health-care utilization. Second, as with all observational studies, residual confounding cannot be excluded despite adjustment for comorbidities, health-care utilization, previous pneumococcal vaccination, influenza vaccination, and other measured characteristics. Notably, the recent Medicare-based evaluation of PCV20 identified evidence of modest residual confounding using prespecified negative-control outcomes. Although our risk-informed propensity score matching approach was designed to further reduce this bias, residual confounding of a similar magnitude cannot be excluded, particularly given the modest vaccine effectiveness estimates observed for pneumonia-related hospitalization. Third, follow-up after PCV20 implementation was limited to two respiratory seasons, precluding evaluation of longer-term effectiveness and durability of protection. Finally, this study was conducted within a single integrated health-care system, and our findings should be confirmed in populations with different pneumococcal serotype distributions, vaccination policies, and health-care settings.

Our study has several important strengths. It included a large, population-based cohort with comprehensive longitudinal electronic health records, enabling capture of vaccination history, hospitalizations, and relevant clinical covariates. The availability of detailed longitudinal data allowed adjustment for previous pneumococcal vaccination, comorbidities, influenza vaccination, and health-care utilization, while our risk-informed propensity score matching approach further reduced confounding inherent to observational vaccine-effectiveness studies. The use of baseline risk stratification also allowed estimation of absolute clinical benefit across clinically relevant risk groups, providing information that is directly relevant to implementation and prioritization.

In conclusion, PCV20 was associated with a reduced risk of pneumonia-related hospitalization among adults aged 65 years or older, but its clinical benefit varied markedly according to baseline pneumonia risk. The majority of older adults were at low predicted risk and experienced minimal absolute benefit, whereas substantially greater benefit was concentrated among medium- and high-risk individuals. These findings demonstrate that population-level estimates of vaccine effectiveness can mask substantial heterogeneity in individual clinical benefit and suggest that incorporating baseline pneumonia risk into vaccination strategies may enable more targeted and potentially better-timed vaccination.

## Data Availability

The individual-level data used in this study are not publicly available because they contain confidential health information from Clalit Health Services electronic health records. Access to the data is restricted to authorized researchers within the secure Clalit Health Services research environment and is subject to institutional and ethical approvals.

