## Supplementary Appendix for "Risk-based vaccination reveals marked heterogeneity in the clinical benefit of PCV20"

### 1. Pneumococcal vaccination uptake

Figure S1 summarizes pneumococcal vaccination uptake during the study period. PPSV23 was the predominant pneumococcal vaccine before the introduction of conjugate vaccination in adults, followed by increasing uptake of PCV13 after its recommendation. Following the introduction of PCV20, uptake increased rapidly during the 2023-2024 and 2024-2025 respiratory seasons, becoming the most frequently administered pneumococcal vaccine among older adults by the end of the study period. Figure S1 also summarizes the distribution of previous pneumococcal vaccination histories, demonstrating that a substantial proportion of participants had received sequential pneumococcal vaccination before PCV20 administration.

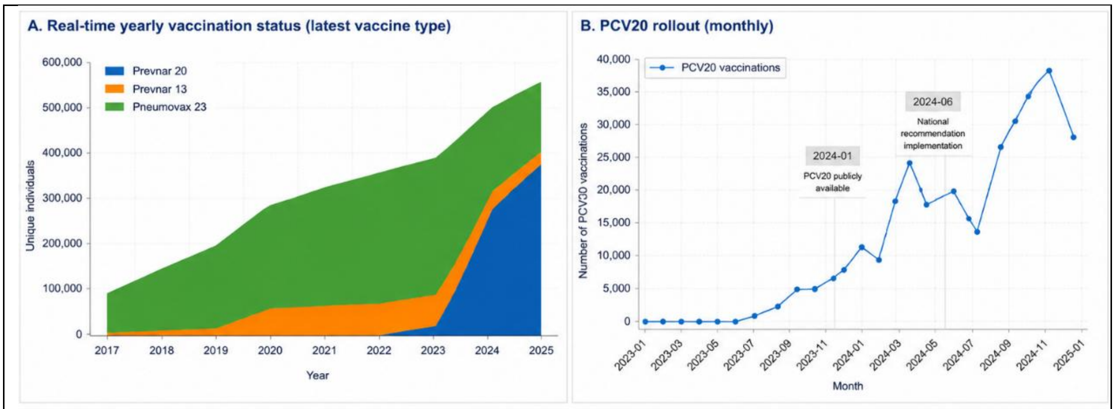

C. Vaccination history combinations (ever received)

| Vaccination history (ever received) | Prevnar 20 (PCV20) | Prevnar 13 (PCV13) | Pneumovax 23 (PPSV23) | Number of individuals n (%) |
|---|---|---|---|---|
| 1. PCV20 only | ✓ | × | × | 163,979 (20.82%) |
| 2. PCV13 only | × | ✓ | × | 20,658 (2.62%) |
| 3. PPSV23 only | × | × | ✓ | 130,674 (16.59%) |
| 4. PCV20 + PCV13 | ✓ | ✓ | × | 35,310 (4.48%) |
| 5. PCV20 + PPSV23 | ✓ | × | ✓ | 153,862 (19.54%) |
| 6. PCV13 + PPSV23 | × | ✓ | ✓ | 11,053 (1.40%) |
| 7. PCV20 + PCV13 + PPSV23 | ✓ | ✓ | ✓ | 23,671 (3.01%) |
| 8. No pneumococcal vaccination | × | × | × | 248,331 (31.53%) |
| Total | | | | 787,538 (100%) |

**Figure S1. Uptake of pneumococcal vaccination during the study period.** (A) Yearly distribution of the most recent pneumococcal vaccine received among adults aged 65 years or older from 2017 to 2025. (B) Monthly rollout of PCV20 vaccinations during the 2023-2024 and 2024-2025 respiratory seasons. (C) Distribution of previous pneumococcal vaccination histories among study participants, including combinations of PCV20, PCV13, and PPSV23.

### 2. Development and validation of the pneumonia risk prediction model

A prediction model was developed to estimate each participant's probability of pneumonia-related hospitalization during the subsequent 12 months. Model development used longitudinal electronic health record data collected between Jan 1, 2017, and Dec 31, 2021. Independent temporal validation was performed using data from Jan 1, 2022, to Dec 31, 2022, before the introduction of PCV20, ensuring complete separation between model development and the vaccine effectiveness analyses.

The outcome was pneumonia-related hospitalization within 12 months after the prediction date. Candidate predictors were prespecified according to clinical relevance and data availability and included demographic characteristics, socioeconomic variables, healthcare utilization, previous pneumonia episodes, previous hospitalizations, influenza vaccination history, medication use, smoking status, comorbidity burden, and chronic medical conditions (Appendix Table S1). An extreme gradient boosting (XGBoost) algorithm was used for model development. Categorical predictors included sex, sector, country of birth, and insurance type, while all other variables were treated as numerical features.

Given that pneumonia-related hospitalization is a relatively infrequent outcome (approximately 1·5-2% annually), class imbalance was addressed using the scale_pos_weight parameter to increase the contribution of positive cases during training. Hyperparameters were optimized using grid search with 20 candidate configurations, conducted on a stratified subsample of 50 000 records to ensure adequate representation of outcome events while maintaining computational efficiency. The hyperparameter search space included n_estimators (300, 500, 700), max_depth (3-6), learning_rate (0.02-0.05), subsample (0.7-1.0), colsample_bytree (0.7-1.0), min_child_weight (1, 3, 5), gamma (0-1.0), reg_alpha (0-1.0), and reg_lambda (3-12). Model selection was based on the area under the precision-recall curve (AUPRC), which is appropriate for imbalanced outcomes.

The final model generated an individualized predicted probability of pneumonia-related hospitalization, which was subsequently used for risk stratification in the primary propensity score matching analysis.

**Model performance**

Model performance was evaluated in the independent 2022 validation cohort. Discrimination was assessed using the area under the precision–recall curve (AUPRC), because pneumonia-related hospitalization was a relatively infrequent outcome. Calibration was assessed by comparing observed and predicted hospitalization rates across the predicted risk spectrum using locally weighted scatterplot smoothing (LOWESS) with bootstrap-derived 95% confidence intervals.

The model demonstrated good discrimination, with an AUPRC of 0·84 (Appendix Figure S2). Calibration was good throughout the predicted risk range, with close agreement between observed and predicted hospitalization rates and only minor deviations among the highest-risk participants, where event numbers were limited (Appendix Figure S2).

Feature importance was assessed using SHapley Additive exPlanations (SHAP). The strongest predictors of pneumonia-related hospitalization were the number of active comorbidities, age, overall comorbidity burden, pulmonary disease, hospitalization during the previous year, insurance type, and neurological disease (Appendix Figure S3A). Previous pneumonia, malignancy, medication use, and healthcare utilization also contributed to model predictions, supporting the clinical plausibility of the model.

**Table S1. Candidate predictors included in the pneumonia risk prediction model.**

| Category | Variables |
|---|---|
| **Demographic characteristics** | Age, sex, sector, country of birth, retirement status |
| **Socioeconomic characteristics** | Insurance type, geographic region, community socioeconomic status |
| **Health-care utilization** | Number of hospitalizations during the previous year; number of hospitalizations during the preceding 1–3 years; number of home-care visits during the previous year; number of home-care visits during the preceding 1–3 years; total number of community diagnoses; any community visit |
| **Previous respiratory disease** | Pneumonia during the previous year; pneumonia during the preceding 1–3 years; home-treated pneumonia during the previous year; home-treated pneumonia during the preceding 1–3 years |

| | |
|---|---|
| **Vaccination history** | Influenza vaccination during the previous three years |
| **Medication use** | Medication groups A, B, and H (see Appendix Methods) |
| **Smoking** | Current smoking status |
| **Chronic medical conditions** | Chronic heart failure, chronic obstructive pulmonary disease, chronic kidney disease, cerebrovascular disease, diabetes mellitus, hypertension, chronic liver disease, malignancy, neurological disease, obesity |
| **Severe immunocompromising conditions*** | Solid organ transplantation, kidney transplantation, dialysis-dependent kidney disease, anatomical or functional asplenia (including splenectomy and sickle cell disease) |
| **Immune-mediated chronic diseases**** | Rheumatoid arthritis, systemic lupus erythematosus, inflammatory bowel disease (Crohn's disease and ulcerative colitis), multiple sclerosis, sarcoidosis, systemic sclerosis, polymyalgia rheumatica |
| **Overall health status** | Presence of any comorbidity, total number of active comorbidities |

* **Severe immunocompromising conditions** correspond to conditions that markedly impair host defence and are largely aligned with ACIP-defined high-risk conditions.

** **Immune-mediated chronic diseases** are inflammatory or autoimmune disorders that may increase infection risk, particularly through immunosuppressive treatment, but are not themselves synonymous with immunodeficiency.

##### Propensity score model

A separate machine learning model was developed to estimate the probability of receiving PCV20 vaccination in the subsequent year. The outcome variable for this model was vaccination during the following year. The model included the same covariates as the pneumonia risk model, with the addition of prior vaccination history variables. The same modelling framework, training strategy, and evaluation approach were applied.

**Table S2 :ICD-9-CM diagnosis codes used to define pneumonia-related outcome.**

| ICD-9-CM code | Diagnosis | Pneumonia-related hospitalization | Outpatient pneumonia |
|---|---|---|---|

| | | | |
|---|---|---|---|
| 382 | Pneumococcal septicaemia | | ✓ |
| 412 | Pneumococcal infection in conditions classified elsewhere, unspecified site | | ✓ |
| 466, 466.0 | Acute bronchitis and bronchiolitis | | ✓ |
| 480-480.9 | Viral pneumonia (including adenovirus, respiratory syncytial virus, parainfluenza virus, SARS-associated coronavirus, and other viral pneumonia) | ✓ | ✓ |
| 481 | Pneumococcal pneumonia | ✓ | ✓ |
| 482-482.9 | Bacterial pneumonia (including *Klebsiella pneumoniae*, *Pseudomonas*, *Haemophilus influenzae*, *Streptococcus*, *Staphylococcus*, and unspecified bacterial pneumonia) | ✓ | ✓ |
| 482.30-482.49 | Streptococcal and staphylococcal pneumonia subtypes | ✓ | ✓ |
| 482.81-482.84 | Pneumonia due to anaerobes, *Escherichia coli*, other Gram-negative bacteria, and Legionnaires' disease | ✓ | ✓ |
| 483.0-483.1 | Pneumonia due to *Mycoplasma pneumoniae* or *Chlamydia* | ✓ | ✓ |
| 484-484.8 | Pneumonia in infectious diseases classified elsewhere (including cytomegalovirus, pertussis, anthrax, aspergillosis, systemic mycoses, and other infectious diseases) | ✓ | ✓ |
| 485 | Bronchopneumonia, organism unspecified | ✓ | ✓ |

| | | | |
|---|---|---|---|
| 486 | Pneumonia, organism unspecified | ✓ | ✓ |
| 487.0 | Influenza with pneumonia | ✓ | ✓ |
| 488.01, 488.11 | Influenza due to avian influenza virus or novel H1N1 influenza virus with pneumonia | ✓ | ✓ |
| 320.1 | Pneumococcal meningitis | | ✓ |

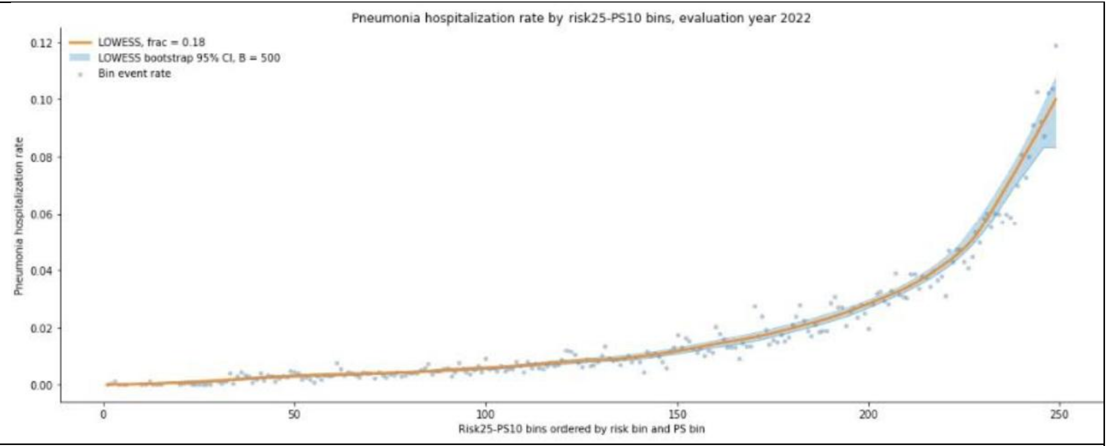

**Figure S2 :Calibration of the pneumonia risk prediction model in the independent 2022 validation cohort.** Observed pneumonia-related hospitalization rates (grey points) are shown across ordered risk–propensity score bins. The solid orange line represents the LOWESS-smoothed observed event rate, and the shaded blue area indicates the bootstrap-derived 95% confidence interval (500 bootstrap iterations). The close agreement between observed and predicted event rates demonstrates good calibration across the predicted risk spectrum.

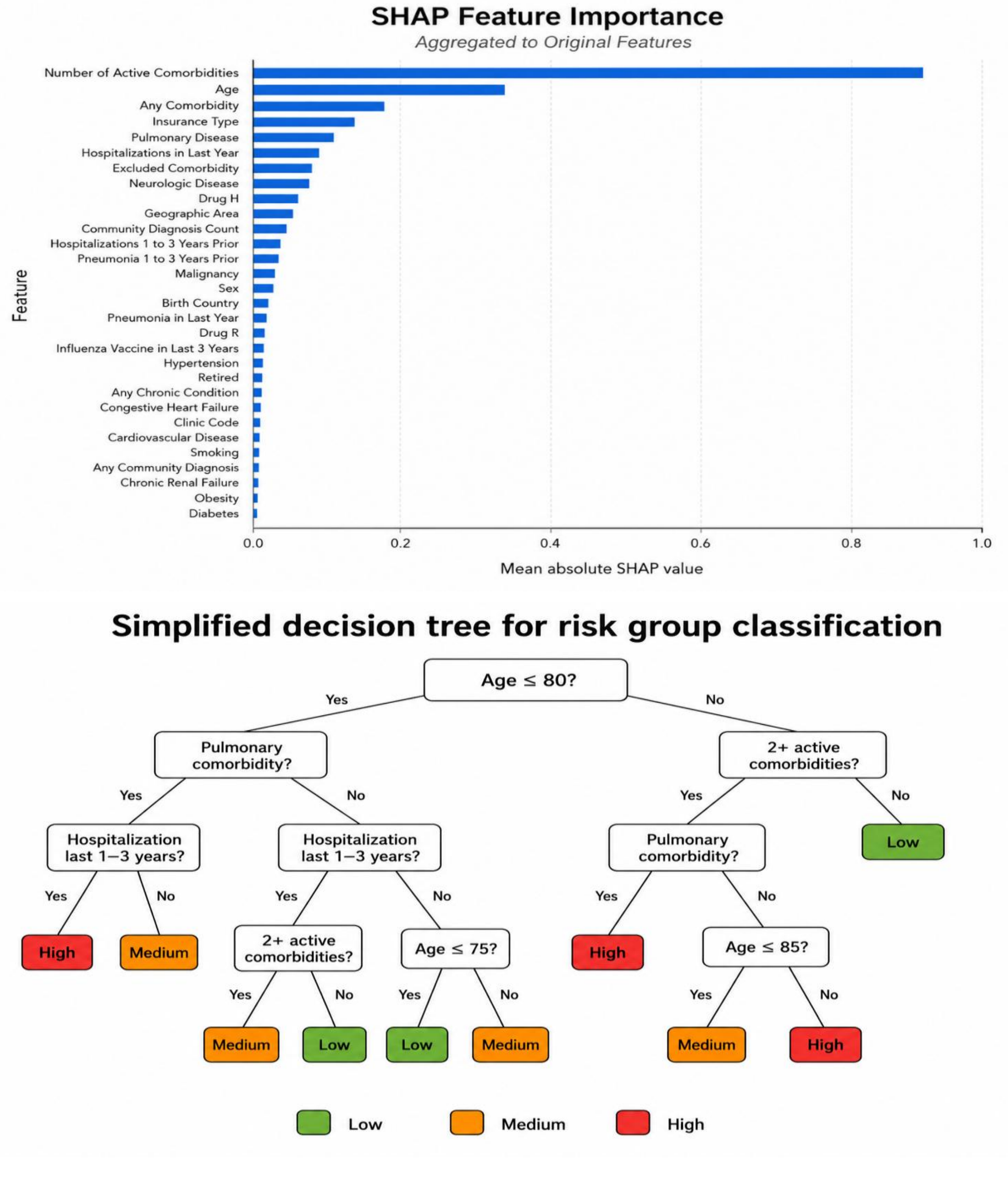

**Figure S3. Interpretation and clinical simplification of the pneumonia risk prediction model. (A)** Mean absolute SHAP values are shown for each predictor in the independent 2022 validation cohort. Larger SHAP values indicate greater influence on the predicted probability of pneumonia-related hospitalization. The number of active comorbidities, age, overall comorbidity burden, pulmonary disease, and previous hospitalization contributed most strongly to model predictions. **(B)** Simplified four-level surrogate decision tree trained to reproduce the low-, medium-, and high-risk categories derived from the XGBoost predictions. The tree uses age, chronic pulmonary disease, hospitalization during the preceding 1-3 years, and the number of active comorbidities to provide a clinically interpretable approximation of the model-derived risk groups. The surrogate tree was intended for descriptive purposes and was not used for matching or vaccine-effectiveness estimation.

##### Clinical simplification of the model-derived risk groups

To improve the clinical interpretability of the model-derived risk categories, we trained a shallow surrogate decision tree to reproduce the low-, medium-, and high-risk group assignments derived from the XGBoost-predicted 12-month risk of pneumonia hospitalization. The tree was restricted to four decision levels and used a small set of clinically interpretable variables, including age, chronic pulmonary disease, hospitalization during the preceding 1-3 years, and the number of active comorbidities.

The surrogate tree was developed as a descriptive approximation of the XGBoost-derived risk groups and informed the development of the simplified clinical scoring approach. It was not used to estimate the original predicted risks, define the 25 matching strata, perform matching, or estimate vaccine effectiveness (Figure S3B).

#### 3. Alternative covariate-based matching analysis

As a sensitivity analysis, we repeated the primary analysis using a conventional covariate-based matching approach rather than the machine learning risk-stratified propensity score framework. This analysis was designed to evaluate the robustness of the primary findings using a simpler prespecified matching scheme based on demographic, sociodemographic, and clinical characteristics. Monthly exact matching was performed on age adjustment category, sex, sociodemographic status, respiratory high-risk status, and other chronic conditions. Age adjustment categories were defined as 65-69, 70-79, and 80 years or older. Sociodemographic status was categorized into tertiles according to the Clalit Health Services socioeconomic index. Respiratory high-risk status was defined as the presence of chronic pulmonary disease or smoking history. Other chronic conditions were defined as the presence of at least one of the following: chronic heart failure, chronic kidney disease, cerebrovascular disease, hypertension, or diabetes mellitus. Variable definitions are summarized in Table S3.

In this alternative analysis, no machine learning risk strata or propensity score strata were used for matching. Instead, in each calendar month, PCV20-vaccinated person-months were matched to eligible unvaccinated control person-months within the same covariate-defined matching cell. Vaccine effectiveness was estimated using the same monthly matching procedure, bootstrap resampling framework, outcome definition, and analytical approach as in the primary analysis.

The results of the alternative covariate-based matching analysis were directionally consistent with the primary risk-stratified propensity score analysis (Figure S4). Estimated vaccine effectiveness against pneumonia hospitalization was 20.3% (95% CI, 8.9 to 29.9) in the 2023-2024 respiratory season and 20.4% (95% CI, 16.6 to 24.2) in the 2024-2025 respiratory season, with a pooled estimate of 20.4% (95% CI, 16.9 to 23.8). Overall, these findings did not materially alter the interpretation of the primary analysis and support the robustness of the main results to an alternative, simpler matching strategy.

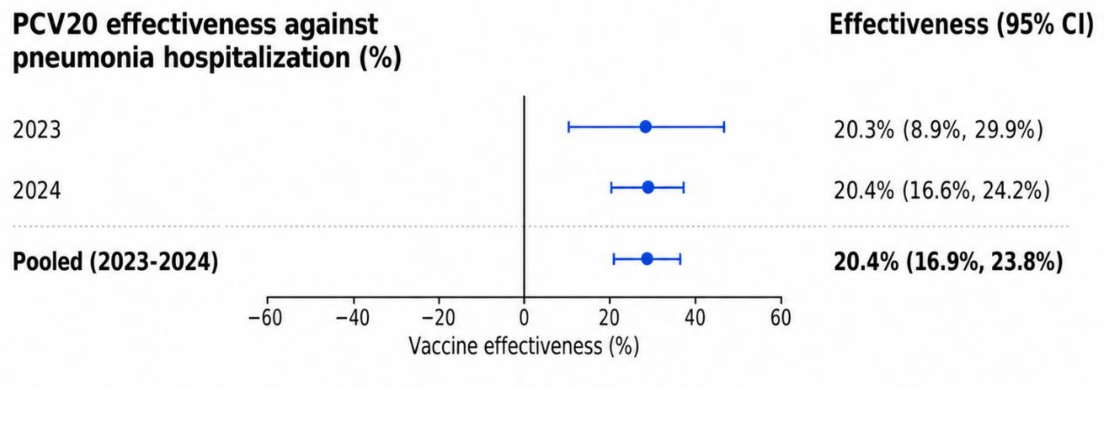

**Figure S4. Sensitivity analysis using conventional covariate-based matching.** Vaccine effectiveness of PCV20 against pneumonia hospitalization, estimated using monthly exact matching on age adjustment category, sex, sociodemographic tertile, respiratory high-risk status, and other chronic conditions.

**Table S3. Covariates used in the alternative covariate-based matching analysis.**

| Matching variable | Definition | Coding |
|---|---|---|
| **Age group** | Age categorized into broad age groups | 65-69 years; 70-79 years; ≥80 years |
| **Sex** | Biological sex | Female; Male |
| **Sociodemographic status** | Clalit Health Services sociodemographic index categorized into tertiles | Lower, middle, and upper tertiles |
| **Respiratory risk factors** | Presence of chronic obstructive pulmonary disease (COPD), asthma, chronic bronchitis, bronchiectasis, or smoking history | Yes / No |
| **Other chronic medical conditions** | Presence of at least one of chronic heart failure, chronic kidney disease, cerebrovascular disease, hypertension, or diabetes mellitus | Yes / No |
